# Clinical phenotypes and outcomes associated with SARS-CoV-2 Omicron variant JN.1 in critically ill COVID-19 patients: a prospective, multicenter cohort study

**DOI:** 10.1101/2024.03.11.24304075

**Authors:** Nicolas de Prost, Etienne Audureau, Antoine Guillon, Lynda Handala, Sébastien Préau, Aurélie Guigon, Fabrice Uhel, Quentin Le Hingrat, Flora Delamaire, Claire Grolhier, Fabienne Tamion, Alice Moisan, Cédric Darreau, Jean Thomin, Damien Contou, Amandine Henry, Thomas Daix, Sébastien Hantz, Clément Saccheri, Valérie Giordanengo, Tài Pham, Amal Chaghouri, Pierre Bay, Jean-Michel Pawlotsky, Slim Fourati, the SEVARVIR investigators

**Author notes:** **Corresponding authors:** Prof. Slim Fourati, Department of Virology, Hôpitaux Universitaires Henri Mondor, Assistance Publique – Hôpitaux de Paris, Créteil, France Prof. N. de PROST Service de Médecine Intensive Réanimation, Hôpital Henri Mondor, Créteil, France. Study Group team members are listed in the Acknowledgments.

## Abstract

A notable increase in severe cases of COVID-19, with significant hospitalizations due to the emergence and spread of JN.1 was observed worldwide in late 2023 and early 2024. During the study period (November 2022-January 2024), 56 JN.1- and 126 XBB-infected patients were prospectively enrolled in 40 French intensive care units. JN.1-infected patients were more likely to be obese (35.7% vs 20.8%; p=0.033) and less frequently immunosuppressed than others (20.4% vs 41.4%; p=0.010). JN.1-infected patients required invasive mechanical ventilation support in 29.1%, 87.5% of them received dexamethasone, 14.5% tocilizumab and none received monoclonal antibodies. Day-28 mortality of JN.1-infected patients was 14.6%.

## BACKGROUND

Following the emergence of the Omicron variant of SARS-CoV-2, several sublineages have co-circulated until the dominance of XBB recombinant variants in early 2023, which were subsequently replaced by a distinct branch of BA.2 named BA.2.86. Compared to XBB and the parental BA.2, the spike protein of BA.2.86 has more than 30 mutations [1]. Initially, BA.2.86 did not dominate other coexisting subvariants until it acquired an additional mutation (i.e., L455S), causing its progeny JN.1 to rapidly increase and become the dominant SARS-CoV-2 variant in several parts of the world. Subsequently, the WHO designated JN.1 as a variant of interest due to its increased transmissibility.

Several *in vitro* studies have shown that JN.1 has phenotypic characteristics that confer enhanced *in vitro* fitness. The L455S substitution in the spike protein enhances the ability of the virus to bind to the angiotensin-converting enzyme 2 receptor. JN.1 also appears to be one of the most immune-evading SARS-CoV-2 variants to date, contributing to its increased transmissibility compared to other Omicron sublineages [2].

Clinical reports from medical institutions indicate that the risk of serious illness due to JN.1 variant infection is low [3]. However, there has been a notable increase in severe cases of COVID-19, with significant hospitalizations due to COVID-19 in late 2023. Importantly, a certain proportion of patients is still admitted to intensive care units (ICUs) for COVID-19-associated acute respiratory failure, but their clinical phenotype and outcomes have changed since the early waves of the pandemic [4,5], and those of patients admitted with severe COVID-19 due to the JN.1 subvariant are currently unknown. This information is critical as it could improve our ability to target individuals who may benefit from more personalized preventive measures, such as frequent vaccination and/or active immunoprophylaxis, as well as tailored therapeutic interventions, including early administration of antivirals in the event of infection.

As part of the SEVARVIR study, we have established a prospective French national multicenter cohort focused on patients admitted to ICUs with COVID-19-associated acute respiratory failure. In this specific substudy, our aim is to comprehensively characterize the clinical presentation and outcomes of patients infected with the emerging JN.1 variant and compare them with those infected with sublineages derived from XBB.

## METHODS

### Study design and patients

The current study is a substudy of the SEVARVIR prospective multicenter observational cohort study. Patients admitted to any of the 40 participating ICUs between November 17, 2022, and January 22, 2024, were eligible for inclusion in the SEVARVIR cohort study (NCT05162508) if they met the following inclusion criteria: age ≥18 years, SARS-CoV-2 infection confirmed by a positive reverse transcriptase-polymerase chain reaction (RT-PCR) in nasopharyngeal swab samples, ICU admission for acute respiratory failure (i.e., peripheral oxygen saturation ≤90% and need for supplemental oxygen or any type of ventilatory support). Patients with SARS-CoV-2 infection but no acute respiratory failure or with a RT-PCR cycle threshold (Ct) value >32 in nasopharyngeal swabs were not included. The study was approved by the Comité de Protection des Personnes Sud-Méditerranée I (N° EudraCT/ID-RCB: 2021-A02914-37). Informed consent was obtained from all patients or their relatives.

Demographics, clinical and laboratory variables were recorded upon ICU admission and during ICU stay. Patients’ frailty was assessed using the Clinical Frailty Scale [6]. The severity of the disease upon ICU admission was assessed using the World Health Organization (WHO) 10-point ordinal scale [7], the sequential organ failure assessment (SOFA) score, and the simplified acute physiology score (SAPS) II score. Acute respiratory distress syndrome (ARDS) was defined according to the Berlin definition [8]. The primary clinical endpoint of the study was day-28 mortality.

### SARS-CoV-2 variant determination

Full-length SARS-CoV-2 genomes from all included patients were sequenced by means of next-generation sequencing. For mutational pattern analysis at the amino acid level, only high-quality sequences, i.e., sequences covering ≥90% of the viral genome and 95% of the spike gene, were considered. Full-length viral genome sequence analysis yielding high coverage have been deposited in Genbank (PP357634 - PP357842).

### Statistical Analysis

Descriptive results are presented as mean±standard deviation [SD] or median (1^st^-3^rd^ quartiles) for continuous variables, and as numbers with percentages for categorical variables. Two-sided p-values <0.05 were considered statistically significant. Unadjusted comparisons between patients infected with two groups of Omicron sublineages (including XBB sublineages, referred to as the “XBB group”, and emerging BA.2.86 sublineages, [parental BA.2.86, JN.1, and JN.3], referred to as the “JN.1 group”) were performed using Chi-squared or Fisher’s exact tests for categorical variables, and ANOVA or Kruskal-Wallis tests for continuous variables, as appropriate.

Analyses were performed with Stata V16.1 statistical software (StataCorp, College Station, TX, USA) and R 4.2.0 (R Foundation for Statistical Computing, Vienna, Austria).

## RESULTS

Between November 17, 2022, and January 22, 2024, 233 patients were admitted to one of the 40 participating ICUs and enrolled in the SEVARVIR cohort study. Of these, 126 patients in the “XBB group” and 56 patients in the “JN.1 group” were included in the analysis.

No statistically significant differences were observed between patients infected with JN.1 and XBB sublineages with respect to age, gender and frequency of comorbidities.

However, patients in the JN.1group were more likely to be obese (n=20/56, 35.7% vs n=26/125, 20.8%; p=0.033), and had a higher median body mass index (26.4 [22.4-33.4] vs 25.0 [21.2-28.7] kg/m^2^; p=0.019). There were also significantly fewer immunosuppressed patients in the JN.1 group than in the XBB group (n=10/49, 20.4% vs n=48/116, 41.4%; p=0.010) (**Table 1**).

**Table 1.**
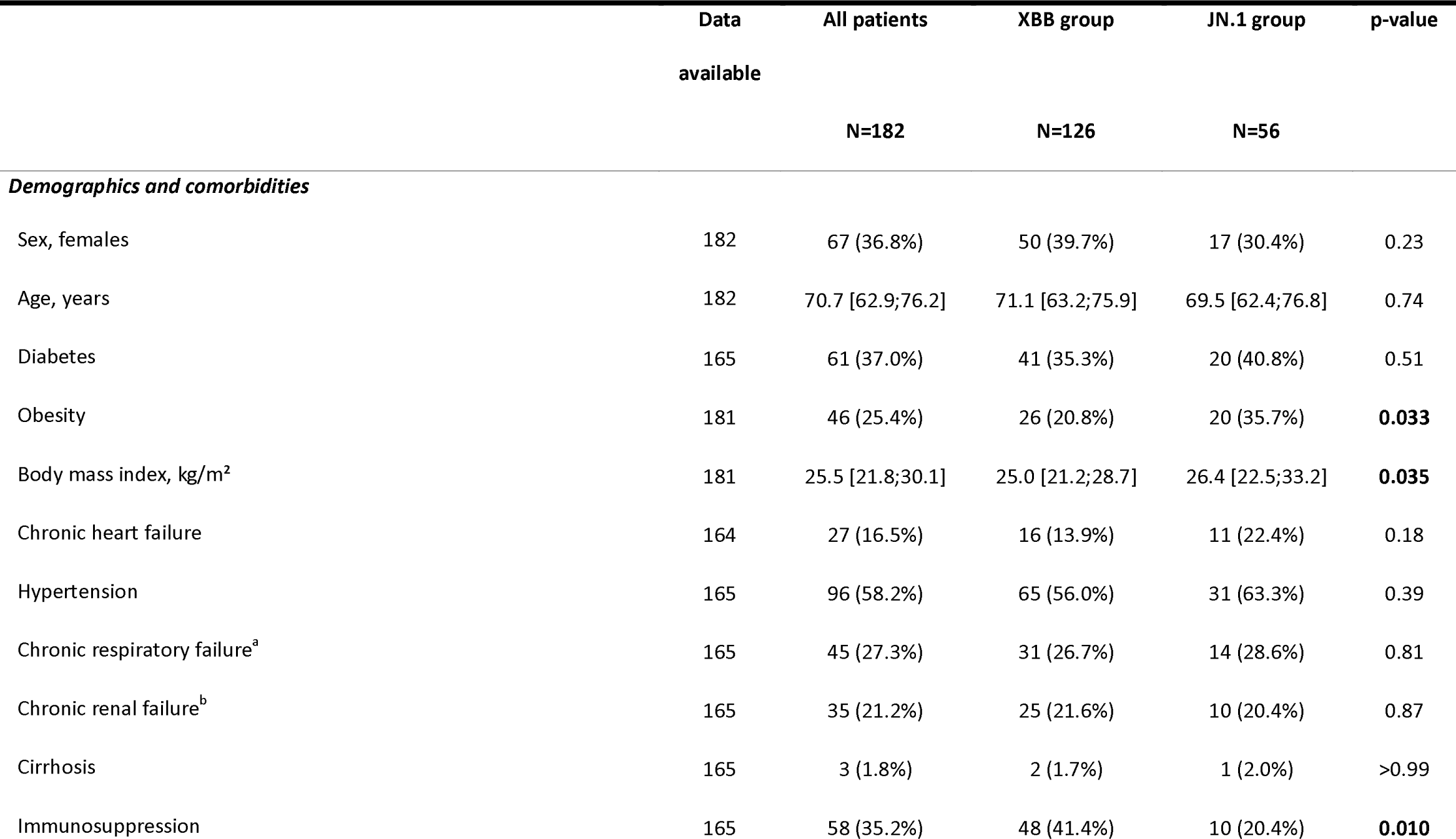
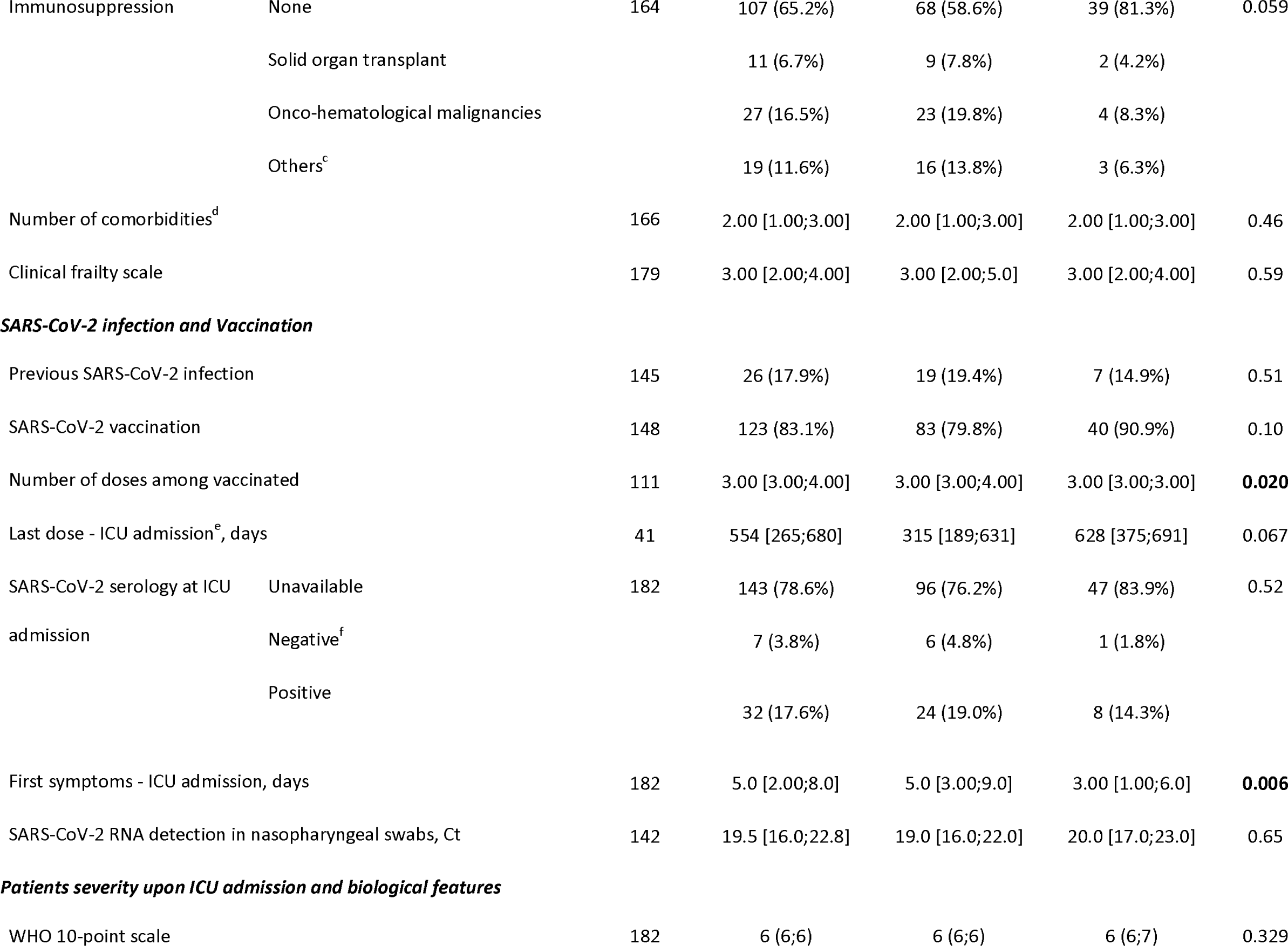
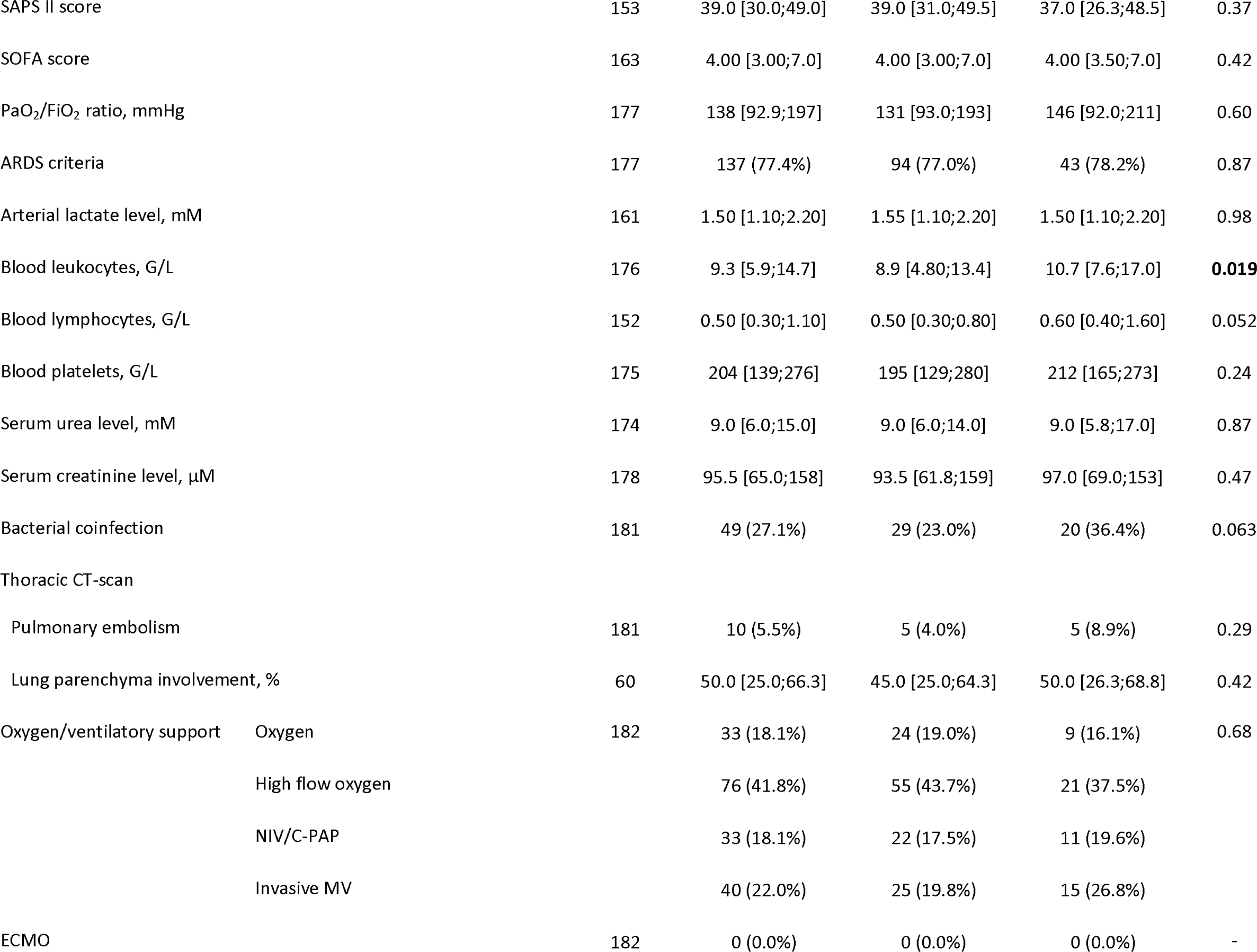
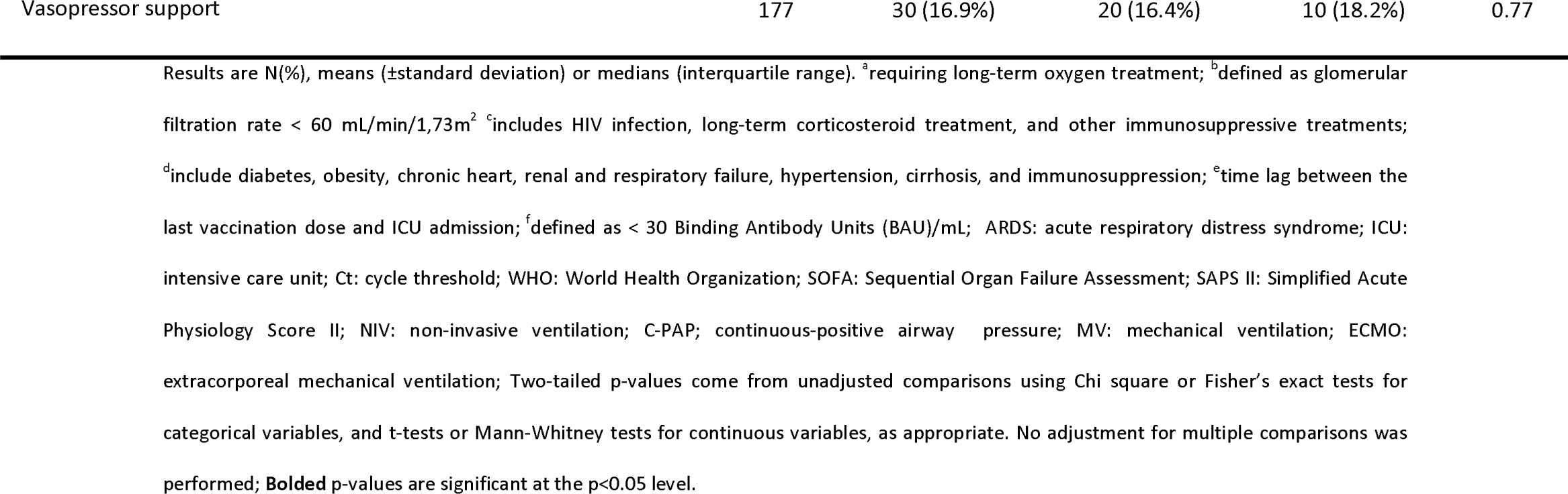
Clinical and biological characteristics of the 182 patients with severe SARS-CoV-2 infection at the time of their intensive care unit admission according to the infecting SARS-CoV-2 “sublineage groups” (XBB vs JN.1 group).

|  | Data<br>available | All patients<br>N=182 | XBB group<br>N=126 | JN.1 group<br>N=56 | p-value |
| --- | --- | --- | --- | --- | --- |
| ***Demographics and comorbidities*** |  |  |  |  |  |
| Sex, females | 182 | 67 (36.8%) | 50 (39.7%) | 17 (30.4%) | 0.23 |
| Age, years | 182 | 70.7 [62.9;76.2] | 71.1 [63.2;75.9] | 69.5 [62.4;76.8] | 0.74 |
| Diabetes | 165 | 61 (37.0%) | 41 (35.3%) | 20 (40.8%) | 0.51 |
| Obesity | 181 | 46 (25.4%) | 26 (20.8%) | 20 (35.7%) | **0.033** |
| Body mass index, kg/m<sup>2</sup> | 181 | 25.5 [21.8;30.1] | 25.0 [21.2;28.7] | 26.4 [22.5;33.2] | **0.035** |
| Chronic heart failure | 164 | 27 (16.5%) | 16 (13.9%) | 11 (22.4%) | 0.18 |
| Hypertension | 165 | 96 (58.2%) | 65 (56.0%) | 31 (63.3%) | 0.39 |
| Chronic respiratory failure<sup>a</sup> | 165 | 45 (27.3%) | 31 (26.7%) | 14 (28.6%) | 0.81 |
| Chronic renal failure<sup>b</sup> | 165 | 35 (21.2%) | 25 (21.6%) | 10 (20.4%) | 0.87 |
| Cirrhosis | 165 | 3 (1.8%) | 2 (1.7%) | 1 (2.0%) | >0.99 |
| Immunosuppression | 165 | 58 (35.2%) | 48 (41.4%) | 10 (20.4%) | **0.010** |

|  |  |  |  |  |  |  |
| --- | --- | --- | --- | --- | --- | --- |
| Immunosuppression | None | 164 | 107 (65.2%) | 68 (58.6%) | 39 (81.3%) | 0.059 |
|  | Solid organ transplant |  | 11 (6.7%) | 9 (7.8%) | 2 (4.2%) |  |
|  | Onco-hematological malignancies |  | 27 (16.5%) | 23 (19.8%) | 4 (8.3%) |  |
|  | Others<sup>c</sup> |  | 19 (11.6%) | 16 (13.8%) | 3 (6.3%) |  |
| Number of comorbidities<sup>d</sup> |  | 166 | 2.00 [1.00;3.00] | 2.00 [1.00;3.00] | 2.00 [1.00;3.00] | 0.46 |
| Clinical frailty scale |  | 179 | 3.00 [2.00;4.00] | 3.00 [2.00;5.0] | 3.00 [2.00;4.00] | 0.59 |
| ***SARS-CoV-2 infection and Vaccination*** |  |  |  |  |  |  |
| Previous SARS-CoV-2 infection |  | 145 | 26 (17.9%) | 19 (19.4%) | 7 (14.9%) | 0.51 |
| SARS-CoV-2 vaccination |  | 148 | 123 (83.1%) | 83 (79.8%) | 40 (90.9%) | 0.10 |
| Number of doses among vaccinated |  | 111 | 3.00 [3.00;4.00] | 3.00 [3.00;4.00] | 3.00 [3.00;3.00] | **0.020** |
| Last dose - ICU admission<sup>e</sup>, days |  | 41 | 554 [265;680] | 315 [189;631] | 628 [375;691] | 0.067 |
| SARS-CoV-2 serology at ICU admission | Unavailable | 182 | 143 (78.6%) | 96 (76.2%) | 47 (83.9%) | 0.52 |
|  | Negative<sup>f</sup> |  | 7 (3.8%) | 6 (4.8%) | 1 (1.8%) |  |
|  | Positive |  | 32 (17.6%) | 24 (19.0%) | 8 (14.3%) |  |
| First symptoms - ICU admission, days |  | 182 | 5.0 [2.00;8.0] | 5.0 [3.00;9.0] | 3.00 [1.00;6.0] | **0.006** |
| SARS-CoV-2 RNA detection in nasopharyngeal swabs, Ct |  | 142 | 19.5 [16.0;22.8] | 19.0 [16.0;22.0] | 20.0 [17.0;23.0] | 0.65 |
| ***Patients severity upon ICU admission and biological features*** |  |  |  |  |  |  |
| WHO 10-point scale |  | 182 | 6 (6;6) | 6 (6;6) | 6 (6;7) | 0.329 |
| SAPS II score |  | 153 | 39.0 [30.0;49.0] | 39.0 [31.0;49.5] | 37.0 [26.3;48.5] | 0.37 |
| SOFA score |  | 163 | 4.00 [3.00;7.0] | 4.00 [3.00;7.0] | 4.00 [3.50;7.0] | 0.42 |
| PaO<sub>2</sub>/FiO<sub>2</sub> ratio, mmHg |  | 177 | 138 [92.9;197] | 131 [93.0;193] | 146 [92.0;211] | 0.60 |
| ARDS criteria |  | 177 | 137 (77.4%) | 94 (77.0%) | 43 (78.2%) | 0.87 |
| Arterial lactate level, mM |  | 161 | 1.50 [1.10;2.20] | 1.55 [1.10;2.20] | 1.50 [1.10;2.20] | 0.98 |
| Blood leukocytes, G/L |  | 176 | 9.3 [5.9;14.7] | 8.9 [4.80;13.4] | 10.7 [7.6;17.0] | **0.019** |
| Blood lymphocytes, G/L |  | 152 | 0.50 [0.30;1.10] | 0.50 [0.30;0.80] | 0.60 [0.40;1.60] | 0.052 |
| Blood platelets, G/L |  | 175 | 204 [139;276] | 195 [129;280] | 212 [165;273] | 0.24 |
| Serum urea level, mM |  | 174 | 9.0 [6.0;15.0] | 9.0 [6.0;14.0] | 9.0 [5.8;17.0] | 0.87 |
| Serum creatinine level, μM |  | 178 | 95.5 [65.0;158] | 93.5 [61.8;159] | 97.0 [69.0;153] | 0.47 |
| Bacterial coinfection |  | 181 | 49 (27.1%) | 29 (23.0%) | 20 (36.4%) | 0.063 |
| Thoracic CT-scan |  |  |  |  |  |  |
| Pulmonary embolism |  | 181 | 10 (5.5%) | 5 (4.0%) | 5 (8.9%) | 0.29 |
| Lung parenchyma involvement, % |  | 60 | 50.0 [25.0;66.3] | 45.0 [25.0;64.3] | 50.0 [26.3;68.8] | 0.42 |
| Oxygen/ventilatory support | Oxygen | 182 | 33 (18.1%) | 24 (19.0%) | 9 (16.1%) | 0.68 |
|  | High flow oxygen |  | 76 (41.8%) | 55 (43.7%) | 21 (37.5%) |  |
|  | NIV/C-PAP |  | 33 (18.1%) | 22 (17.5%) | 11 (19.6%) |  |
|  | Invasive MV |  | 40 (22.0%) | 25 (19.8%) | 15 (26.8%) |  |
| ECMO |  | 182 | 0 (0.0%) | 0 (0.0%) | 0 (0.0%) | - |
| Vasopressor support | 177 | 30 (16.9%) | 20 (16.4%) | 10 (18.2%) | 0.77 |

Results are N(%), means ( ± standard deviation) or medians (interquartile range). <sup>a</sup>requiring long-term oxygen treatment; <sup>b</sup>defined as glomerular filtration rate < 60 mL/min/1.73m<sup>2</sup> <sup>c</sup>includes HIV infection, long-term corticosteroid treatment, and other immunosuppressive treatments; <sup>d</sup>include diabetes, obesity, chronic heart, renal and respiratory failure, hypertension, cirrhosis, and immunosuppression; <sup>e</sup>time lag between the last vaccination dose and ICU admission; <sup>f</sup>defined as < 30 Binding Antibody Units (BAU)/mL; ARDS: acute respiratory distress syndrome; ICU: intensive care unit; Ct: cycle threshold; WHO: World Health Organization; SOFA: Sequential Organ Failure Assessment; SAPS II: Simplified Acute Physiology Score II; NIV: non-invasive ventilation; C-PAP: continuous-positive airway pressure; MV: mechanical ventilation; ECMO: extracorporeal mechanical ventilation; Two-tailed p-values come from unadjusted comparisons using Chi square or Fisher's exact tests for categorical variables, and t-tests or Mann-Whitney tests for continuous variables, as appropriate. No adjustment for multiple comparisons was performed; **Bolded** p-values are significant at the p<0.05 level.

The proportion of patients who had received at least one dose of SARS-CoV-2 vaccine did not differ significantly between groups, although the median number of doses received was significantly higher in patients from the XBB group (3 [3–4] vs 3 [3–3]; p=0.019). The median time from onset of first symptoms to ICU admission was significantly shorter in the JN.1 group than in the XBB group (3 [1–6] vs 5 [3–9]; p=0.006). Other variables related to SARS-CoV-2 virological characteristics, including median viral level in the upper respiratory tract measured by cycle threshold in RT-PCR and prevalence of positive SARS-CoV-2 anti-S antibodies at ICU admission, did not differ significantly between groups (**Table 1**).

There was no significant difference between the two groups in the severity of illness at ICU admission, as reflected by the SOFA and SAPS II scores and the WHO 10-point ordinal scale (**Table 1**). Invasive mechanical ventilation support was required in 22.0% (n=40/182) of patients within 24 hours of ICU admission, with no significant difference between groups. No patient required extracorporeal membrane oxygenation (ECMO) support on ICU admission.

Respiratory failure was eventually attributed to SARS-CoV-2 pneumonia without bacterial co-infection in about half of cases in both JN.1 and XBB groups (**Table 2**).

**Table 2.**
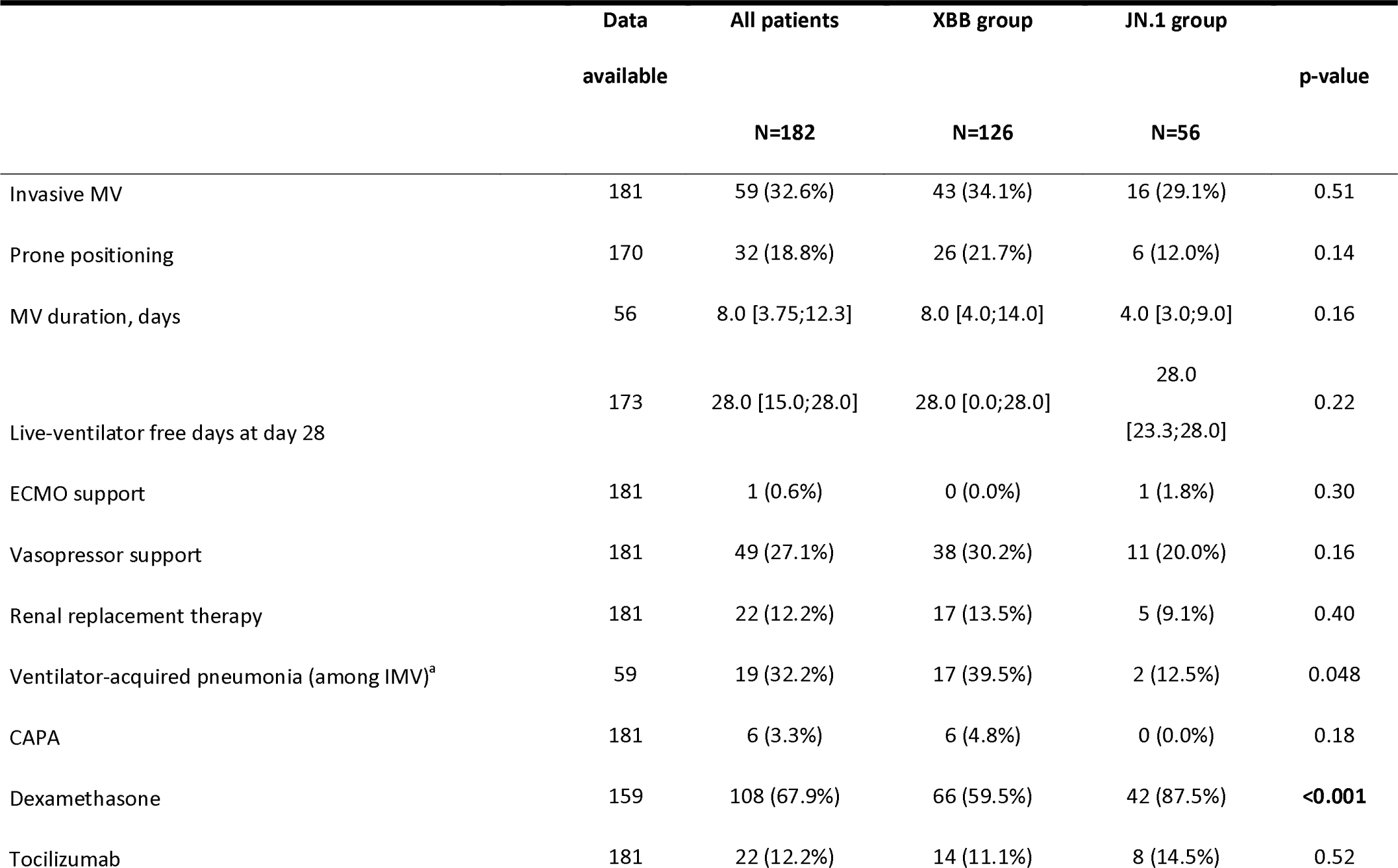
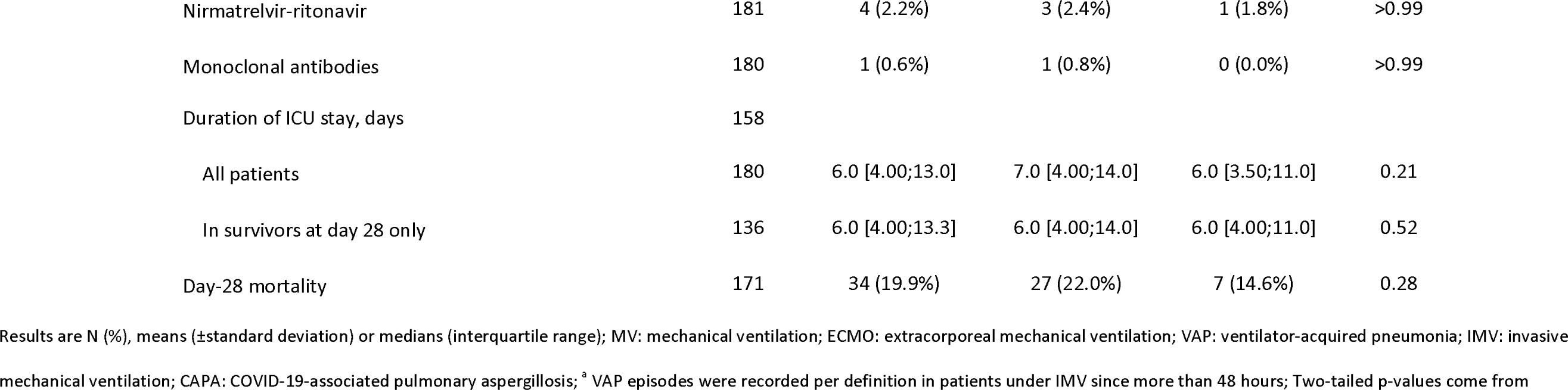
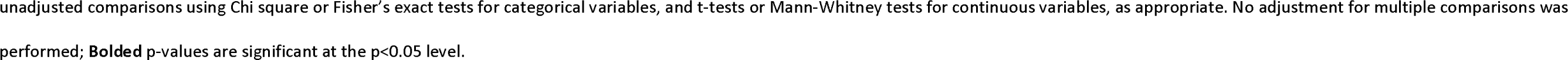
Intensive care management and outcomes of patients with severe SARS-CoV-2 infection (n=182) during their intensive care unit stay according to the SARS-CoV-2 infecting “sublineage groups” (XBB vs JN.1 group).

During the ICU stay, 32.6% (n=59/181) of patients required invasive mechanical ventilation, with no significant differences between the subgroups. There was also no significant difference between groups regarding the need for other organ support (**Table 2**). Day-28 mortality and ICU length of stay were not significantly different between groups. Regarding COVID-19 management, JN.1-infected patients were treated with dexamethasone significantly more often than their XBB counterparts (n=42/48, 87.5% vs n=66/111, 59.5%; p<0.001). No significant differences between groups were observed in the use of other treatments, including anti-IL-6 antagonists, convalescent plasma and antivirals (**Table 2**).

## DISCUSSION

The current study is the first to describe the clinical phenotype associated with the newly emerging Omicron sublineage JN.1 in patients with severe COVID-19 requiring ICU admission. Our data provide reassuring evidence that this emerging sublineage does not cause more severe outcomes than XBB variants that emerged and spread earlier in the population. We observed unexpected phenotypic differences, with more frequent obesity and less frequent immunosuppression in patients infected with JN.1, as compared to those infected with XBB sublineages.

Recent epidemiologic data have confirmed the increased transmissibility of JN.1. Its proportion of the circulating variants in the US had increased to more than 90% according to nowcast estimates from the US Centers for Disease Control and Prevention (CDC) [9]. In France, JN.1 represented more than 90% of circulating variants, according to the *Santé Publique France* report of January 31^st^, 2024 [10]. In this context, and given the surge in COVID-19 cases during the winter of 2024 [3], obtaining clinical data reporting the clinical phenotype and lethality of patients infected with this subvariant as compared with the previous ones is crucial to inform public health authorities and clinicians managing these patients. Our data provide reassuring evidence regarding the severity of disease associated with JN.1 infection, showing not only a non-significant difference in day 28 mortality compared to patients infected with XBB, but also no significant differences in other outcomes, including the need for invasive mechanical ventilation and length of stay in the ICU.

Patients infected with sublineage JN.1 were more likely to be obese and less likely to be immunosuppressed than those infected with XBB in our study. Such a finding was unexpected because immunosuppression has been reported to be the most common comorbidity in COVID-19 patients infected with the Omicron variant since the “ancestral” BA.1 Omicron sublineage [4,11], occuring in almost 50% of cases, and may reflect an inherently less pathogenic variant as reported in a hamster model [12]. On the other hand, the higher prevalence of obesity, a previously reported risk factor for severity with previous SARS-CoV-2 variants, including the ancestral variant, is consistent with previous data reporting obesity as a risk factor for severity [13]. These findings may have important implications for the updated use of pre-exposure monoclonal antibodies use [14] as well as COVID-19 vaccination recommendations. Initial estimates of the updated XBB.1.5 COVID-19 vaccine showed sustained vaccine efficacy against symptomatic JN.1 lineage infection [15]. In terms of ICU management, patients in the JN.1 group received dexamethasone more frequently than their counterparts in the XBB group, possibly because they were less likely to be immunosuppressed. Other aspects of treatment did not differ.

Our study has limitations, including a limited sample size in the JN.1 group, which limitis our statistical power to perform subgroup analyses and adjust for confounding variables.

In conclusion, critically-ill patients with Omicron JN.1 infection showed a different clinical phenotype than patients infected with the earlier XBB sublineage, including more frequent obesity and less immunosuppression. Compared with XBB, JN.1 infection was not associated with significantly different day-28 mortality.

## FOOTNOTE PAGE

### FUNDING STATEMENT

This work was supported by the EMERGEN consortium— ANRS Maladies Infectieuses Emergentes (ANRS0153). This study has been labeled as a National Research Priority by the National Orientation Committee for Therapeutic Trials and other researches on Covid-19 (CAPNET). The investigators would like to acknowledge ANRS | Emerging infectious diseases for their scientific support, the French Ministry of Health and Prevention and the French Ministry of Higher Education, Research and Innovation for their funding and support.

The current work was not presented previously to a meeting.

### CORRESPONDING AUTHOR

Correspondence and requests for reprints should be addressed to Prof Fourati: Department of Virology, Hôpitaux Universitaires Henri Mondor, Assistance Publique – Hôpitaux de Paris, Créteil, France; Tel: +33 1 45 17 81 45

## Data Availability

All data produced in the present study are available upon reasonable request to the authors

## ACKNOWLEDGMENTS

The authors would like to thank all staff involved in the study, Dr Pierre-André Natella, Ms. Nolwenn Bombenger for taking care of regulatory aspects, Mr. Léo Graca for taking care of data management, Mr. Mohamed Ader for clinical data abstraction, the nurses and physicians who took care of the patients, the laboratory staff who took care of virological samples and the patients and their family for agreeing to participate in the study. Assistance Publique – Hôpitaux de Paris is the sponsor of the study.

The authors would like to thank the **SEVARVIR investigators**: Henri Mondor, Créteil, Medical ICU: Nicolas DE PROST, Pierre BAY, Keyvan RAZAZI, Armand MEKONTSO DESSAP; Henri Mondor, Créteil, Surgical ICU: Raphaël BELLAÏCHE, Lucile PICARD; Henri Mondor Créteil, Virology: Slim FOURATI; Alexandre SOULIER; Mélissa N’DEBI; Sarah SENG; Christophe RODRIGUEZ; Jean-Michel PAWLOTSKY; Cochin, Paris, Medical ICU: Frédéric PENE; Cochin, Paris, Virology: Anne-Sophie L’HONNEUR; Saint-Louis, Paris, Medical ICU: Adrien JOSEPH, Elie AZOULAY; Saint-Louis, Paris, Virology: Maud SALMONA; Marie-Laure CHAIX; Pitié-Salpêtrière, Paris, Medical and cardiac ICU: Charles-Edouard LUYT, David LEVY; Pitié-Salpêtrière, Paris, Medical ICU: Julien MAYAUX; Pitié-Salpêtrière, Paris, Virology: Stéphane MAROT, Saint-Antoine, Paris, Medical ICU: Juliette BERNIER; Maxime GASPERMENT, Tomas URBINA, Hafid AIT-OUFELLA, Eric MAURY; Saint-Antoine, Paris, Virology: Laurence MORAND-JOUBERT; Djeneba BOCAR FOFANA; Bichat, Paris, Medical ICU: Jean-François TIMSIT; Bichat, Paris, Virology: Diane DESCAMPS, Quentin LE HINGRAT; Tenon, Paris, Medical and Surgical ICU: Guillaume VOIRIOT, Nina DE MONTMOLLIN, Mathieu TURPIN; Avicenne, Bobigny, Medical and Surgical ICU: Stéphane GAUDRY; Avicenne, Bobigny, Virology: Ségolène BRICHLER; Louis Mourier, Colombes, Medical and Surgical ICU: Fabrice Uhel, Damien Roux; Bicêtre, Le Kremlin-Bicêtre, Medical ICU: Tài Olivier PHAM; Bicêtre, Le Kremlin-Bicêtre, Virology: Amal CHAGHOURI; Raymond Poincaré, Garches, Medical ICU: Nicholas Heming, Djillali Annane; Ambroise Paré, Boulogne, Medical and Surgical ICU: Sylvie Meireles, Antoine Vieillard-Baron; Ambroise Paré, Boulogne, Virology: Elyanne GAULT; Hôpital Marc Jacquet, Melun, Medical ICU: Sébastien Jochmans; Hôpital Marc Jacquet, Melun, Microbiology: Aurélia PITSCH; CH Sud Francilien, Jossigny, ICU: Guillaume CHEVREL, Céline CLERGUE; CH Sud Francilien, Jossigny, Microbiology: Kubab SABAH; CH Victor Dupouy, Argenteuil, ICU: Damien CONTOU; CH Victor Dupouy, Argenteuil, Microbiology: Laurence COURDAVAULT VAGH WEINMANN; Saint-Camille, Bry-sur-Marne, Polyvalent ICU: Malo EMERY; Saint-Camille, Bry-sur-Marne, Microbiology: Claudio GARCIA-SANCHEZ; CHU de Strasbourg, Medical ICU: Ferhat MEZIANI; Louis-Marie JANDEAUX; CHU de Strasbourg, Virology: Samira FAFI-KREMER; Elodie LAUGEL; CHU de Lille, Medical ICU: Sébastien PREAU, Raphaël Favory; CHU de Lille, Virology, Aurélie GUIGNON; CHRU de Nancy, Hôpitaux de Brabois, Medical ICU: Antoine KIMMOUN; CHRU de Nancy, Hôpitaux de Brabois, Virology: Evelyne SCHVOERER, Cédric HARTARD; Antoine Béclère, Clamart, General ICU: Charles DAMOISEL; Hôpital Européen Georges Pompidou, Paris, Medical ICU: Nicolas BRECHOT; Hôpital Européen Georges Pompidou, Paris, Virology: Hélène PÉRÉ; CHU Tours, Medical ICU: Antoine GUILLON; CHU Tours, Virology: Lynda Handala; CHU d’Angers, Medical ICU: François BELONCLE; CHU d’Angers, Virology: Francoise LUNEL FABIANI; CHU de Poitiers, Medical ICU: Rémi COUDROY, Arnaud W. THILLE, François ARRIVE, Sylvain LE PAPE, Laura MARCHASSON; CHU de Poitiers, Virology: Luc DEROCHE, Nicolas LEVEQUE; CHU de Rennes, Medical ICU: Jean-Marc Tadié, Flora DELAMAIRE; CHU de Rennes, Virology: Vincent THIBAUT; Claire GROLHIER; CH de Lorient, Medical ICU: Béatrice LA COMBE; CH de Lorient, Microbiology: Séverine HAOUISEE; CHU de Bordeaux, Medical ICU: Alexandre BOYER; CHU de Bordeaux, Virology: Sonia BURREL; CHU de Rouen, Medical ICU: Fabienne TAMION; Gaetan BEDUNEAU, Christophe GIRAULT, Maximillien GRALL, Dorothée CARPENTIER; CHU de Rouen, Virology: Alice, MOISAN; Jean-Christophe PLANTIER; CHU de Nantes, Medical ICU: Emmanuel CANET; CHU de Nantes, Virology: Audrey, RODALLEC, Berthe Marie IMBERT; CHU de Nice, Medical ICU: Clément SACCHERI; CHU de Nice, Virology: Valérie GIORDANENGO; Marseille Hôpital Nord, Medical ICU: Sami HRAEICH; Marseille Hôpital Nord, IHU Méditerranée: Pierre-Edouard FOURNIER, Philippe COLSON; CH Le Mans, General ICU: Cédric Darreau; CH Le Mans, Microbiology: Jean THOMIN; CHU de Grenoble, Medical ICU: Anaïs DARTEVEL; CHU de Grenoble, Virology: Sylvie LARRAT; CHU de Saint-Etienne, Medical ICU: Guillaume THIERY; CHU de Saint-Etienne, Virology: Sylvie PILLET; CHU de Montpellier, Medical ICU: Kada KLOUCHE; CHU de Montpellier, Virology: Edouard TUAILLON; CHU de Brest, Medical ICU: Cécile AUBRON; CHU de Brest, Virology: Adissa TRAN, Sophie VALLET; CHU de Dijon, Medical ICU: Pierre-Emmanuel CHARLES; CHU de Dijon, Virology: Alexis DE ROUGEMONT; CHU de Clermont-Ferrand, Medical ICU: Bertrand SOUWEINE; CHU de Clermont-Ferrand, Virology: Cecile HENQUELL; Audrey MIRAND; CHU de Reims, Medical ICU: Bruno MOURVILLIER; CHU de Reims, Virology: Laurent ANDREOLETTI, Clément LIER; CHU de Caen, Medical ICU: Damien DU CHEYRON; CHU de Caen, Virology: Nefert CANDACE DOSSOU; Astrid VABRET; CHU de Besançon, Medical ICU: Gaël PITON; CHU de Besançon, Virology: Quentin LEPILLER; CHU de Limoges, Medical ICU: Thomas DAIX; CHU de Limoges, Virology: Sébastien HANTZ, Sylvie ROGER

## Author’s contributions

N.D.P., E.A., J.M.P., and S.F., designed the study and obtained funding; E.A. performed statistical analyses; N.D.P., A.G., S.P., F.U., F.D., F.T., C.D., D.C., T.D., C.S., T.P., P.B., included the patient and were responsible for clinical data collection; L.H., A.G., Q.L.H., V.T., A.M., J.T., A.H., S.H., V.G., A.C., and S.F. were responsible of the management of virological samples; J.-M.P., and S.F. were responsible of virological analyses; N.D.P., E.A., and S.F. wrote the first draft of the article; All authors revised and approved the article. The corresponding author attests that all listed authors meet authorship criteria and that no others meeting the criteria have been omitted. N.D.P. and S.F. are the guarantors.

